# Medication errors in Intensive Care Unit: Assessment of Knowledge among Critical Care Nurses and implementation of a simple strategy to reduce errors

**DOI:** 10.1101/2025.07.07.25331001

**Authors:** Sabin Koirala, Agnimshwor Dahal, Rahul Parajuli, Prashant Acharya, Rebicca Pradhan, Pratibha Paudel

**Affiliations:** Hospital for Advanced Medicine and Surgery (HAMS), Kathmandu, 44600, Nepal; Nepal Intensive Care Research Foundation (NICRF), Kathmandu, 44600, Nepal; Hospital for Advanced Medicine and Surgery, Kathmandu, 44600, Nepal; Kathmandu University (KU), Dhulikhel, 45210, Nepal

**Keywords:** Knowledge, Perception, Behavior, Medication Error, Critical Care Nurses

## Abstract

**Background:** Medication errors are frequent but preventable. Critically ill patients may suffer, on average, 1.7 medical errors each day & some of which may be life-threatening. Such errors can be life-threatening because of the severity of the patients and comorbidities they have.

**Objectives:** To assess the knowledge about medication error and safety among critical care nurses and assess the effect of implementation of focused intervention sessions to improve medication safety.

**Methods:** This is a quantitative cross-sectional study, including all nurses working in critical care areas of a tertiary level hospital. A validated questionnaire was used to collect data on Knowledge, Perception & Behavior of nurses on medication errors and patient safety. Demographic data were analyzed using descriptive statistics. All data were analyzed via SPSS using McNemar’s test.

**Results:** Our study found high baseline compliance with protocol-driven safety practices. There was a significant difference in nurses’ knowledge after education intervention in the use of CPOE reducing medication errors (p 0.00) and alarm noises and ward emergencies increase error risk (p 0.001). Significant differences were noted in perception of the effects of protocols/ guidelines on ensuring proper management of therapeutic processes (p 0.014). The behavioral findings demonstrate near-perfect baseline compliance across all safety practices (98.4-100%), leaving minimal room for measurable improvement.

**Conclusion:** This study highlights significant impact of targeted education on improving critical care nurses’ understanding of computerized order entry and alarm-related risks, reinforcing protocol adherence. Our findings confirm mature safety cultures, and focused interventions effective than broad retraining efforts.

**Executive Summary:** Medication errors are frequent; they can be serious, however, are also preventable. Actually, health care delivery is not perfect and errors are very common. In intensive care units (ICU), critically ill patients may suffer, on average, 1.7 medical errors each day and some of which may be life-threatening. Medication errors can happen anywhere in the hospital; however, those happening in intensive care units can be life-threatening because of the severity of the patients and various comorbidities they have. Almost one-fifth (19%) of the errors in ICU are life-threatening and almost half (42%) cause the addition of other life-sustaining supports, adding emotional, psychological and financial burdens to the patient and family. Although medication errors are increasingly accepted as a significant patient safety issue, research and studies on this topic is limited in low and middle income countries (LMICs), where healthcare systems face various restrictions. There has been minimal research on the knowledge, attitudes, and behaviors of critical care nurses concerning medication safety, especially in the administration of intravenous medications. This study aims to address the gap by evaluating ICU nurses’ comprehension of medication errors, their compliance with safety procedures, and the effect of focused interventions on enhancing safe medication practices.

The objective of this study is to evaluate the nurses’ understanding of medication errors and patient safety. We planned to assess knowledge, perception and behavior of nurses regarding patient safety. After that, we planned to implement and analyze the impact of a focused intervention session, mainly informative meetings with each participant. Thus, we conducted a quantitative, quasi experimental, cross-sectional study with pretest - posttest design. For this, we selected nurses working in critical care settings in a tertiary hospital, namely in the ICU, CCU, HDU and post-operative ward. A validated questionnaire covering key variables - knowledge, perception, behavior and demographic details was used. Post-intervention sessions were conducted 1 week after data collection pertaining to explanation and discussion of correct answers to enhance knowledge. All the responses pretest and posttest were coded for McNemar’s test to assess statistical significance. Data were analyzed using SPSS, with confidence level 95%.

Our study found high baseline compliance with protocol-driven safety practices. There was a significant difference in nurses’ knowledge after education intervention in the use of CPOE reducing medication errors (p 0.00) and alarm noises and ward emergencies increase error risk (p 0.001). Statistically significant differences were noted in participants’ perception of the effects of protocols/ guidelines/ procedures on ensuring proper management of therapeutic processes (p 0.014). The behavioral findings demonstrate near-perfect baseline compliance across all safety practices (98.4-100%), leaving minimal room for measurable improvement through intervention.

This study highlights the significant impact of targeted education on improving critical care nurses’ understanding of computerized order entry and alarm-related risks, reinforcing protocol adherence where initial agreement was incomplete. Despite these improvements, the near-perfect baseline compliance with core medication safety behaviors and strong pre-existing safety perceptions indicate that most fundamental practices are already well institutionalized within the clinical setting. These findings suggest that in mature safety cultures, focused interventions are more effective than broad retraining efforts, as educational programs yield greater success when directed at specific areas requiring enhancement rather than reinforcing well-established norms. We acknowledge there is a need for further research in developing country settings to establish broader trends.

## 1. Introduction

Health care delivery is not perfect and errors are very common. Medication errors are frequent and can be serious, however, are also preventable.. In intensive care units (ICUs), critically ill patients may suffer, 1.7 medical errors each day on average, some of which may be life threatening.^1,2^ The overall medication process involves many steps which can broadly be categorized into 5 stages: prescription, transcription, preparation, dispensation and administration.^3^ Errors can happen in any of these stages, but most commonly they occur during the administration stage (53%), followed by prescription (17%), preparation (14%) and transcription (11%) stages.^4^ The medical errors can be further classified as errors of omission and errors of commission. An error of omission is a failure to perform an appropriate action and an error of commission is defined as performing an inappropriate action.^2^

Medication errors can happen anywhere in the hospital but those happening in intensive care units can be life-threatening because of the severity of the patients and various comorbidities they have. Almost one-fifth (19%) of the errors in ICU are life-threatening and almost half (42%) cause addition of other life-sustaining supports, adding emotional, psychological and financial burden to the patient and family.^5^ The total incidence of medication errors is very difficult to ascertain, largely because of underreporting. There can be many risk factors for medication errors in ICU, with patient illness severity being the strongest predictor of adverse reaction from a medication error.^6,7^

Medication errors are preventable. Different strategies implemented in various hospitals have shown to decrease such errors. Improving the safety of the medication process is the safest and most efficient means of improving patient safety.^8^ This can be accomplished by optimizing the safety of the medication process, eliminating situational risk factors, and providing strategies to both intercept errors and mitigate their consequences. Different strategies that have proved to be successful include: medication standardization,^9,10^ implementation of computerized physicians order entry (CPOE),^11^ medication reconciliation,^12^ using bar-code technology,^13^ and computerized intravenous infusion devices.^3^

Although medication errors are increasingly accepted as a significant patient safety issue, research and studies on this topic is limited in low and middle-income countries (LMICs), where healthcare systems face various restrictions. There has been minimal research on the knowledge, attitudes, and behaviors of critical care nurses concerning medication safety, especially in the administration of intravenous medications. This study aims to address the gap by evaluating ICU nurses’ comprehension of medication errors, their compliance with safety procedures, and the effect of focused interventions on enhancing safe medication practices.

## 2. Methodology

### 2.1. Design

This study was a quantitative, quasi-experimental investigation with a pretest-posttest design to assess critical care nurses’ knowledge, perception, and behaviors regarding medication safety.

### 2.2. Sample and Settings

All critical care nurses working in ICU, HDU, CCU, and postoperative wards in a tertiary-level hospital were included. A convenience sampling method was used for participant selection.

### 2.3. Data Collection

Participants were interviewed using a validated questionnaire developed by Di Muzio et al., which had high reliability (Cronbach’s alpha = 0.776).^14^ The questionnaire included variables such as age, gender, qualification, years of experience, training history, knowledge of medication errors, attitudes toward medication errors, behavior to prevent medication errors, and post-intervention improvements in knowledge, attitude, and behavior. Responses were recorded on a three-level Likert scale (Agree, Disagree, Neutral). The pre-test data was collected from October 15 to November 15, 2024.

Subsequently, a focused intervention session was then conducted from November 16 to November 30, 2024, either in-person to educate participants on strategies for preventing medication errors. The intervention consisted of detailed discussions of the questionnaire items, explaining the correct answers. One week later, a post-intervention test was administered using the same questionnaire to assess knowledge improvements from December 1 to December 31, 2024.

### 2.4. Ethical Considerations

Informed consent was obtained from all participants before data collection. A consent form was attached to the questionnaire to ensure voluntary participation and ethical compliance. Before data collection was considered, staff in all the departments were informed in person and via social media texting apps like Viber and WhatsApp regarding the voluntary nature of the study. In addition, Google Forms had detailed description of the study consisting of Confidentiality and Voluntary Nature of the study as separate topics in the description. Only those who consent were proceeded to the questionnaires and those who do not were directed to the end page of the study.

### 2.5. Measures

The study variables included demographic characteristics (age, gender, qualification, years of experience, and training history) as well as knowledge, perception, and behaviors related to medication errors. The Likert scale responses were converted into binary values for statistical analysis, where “Agree” was coded as 1, and “Disagree” and “Neutral” were coded as 0—as the latter reflected the lack of knowledge or indecision.

### 2.6. Data Analysis

All the collected data were anonymized, and no identifiers were used. Data were only available to the research team. Data collected were entered into Google Forms and exported to MS Excel/SPSS version 26 for analysis. Forms with more than 10% missing data were excluded, and missing responses were recorded as ‘unknown.’ Descriptive statistics were used to analyze demographic data (frequencies and percentages).

Pretest data on knowledge, perception, and behavior were collected, followed by the intervention. The same questionnaire was then used in the post-test phase with the same sample. McNemar’s test was applied to determine changes in responses. Since McNemar’s test requires dichotomous data, responses were converted accordingly. A significance level of 0.05 with a 95% confidence interval was set for statistical analysis.

## 3. Results

### 3.1 Demographic details

The pre-test included 86 participants, while the post-test had 70 participants. To ensure accurate one-to-one comparisons using the McNemar test, we selected the first 70 participants from the pre-test and all 70 participants from the post-test. The remaining 16 participants from the pre-test were excluded from the analysis for comparability.

For demographic details, we are using the pre-test group which includes all the post-test group participants as well (Table 1). All participants were females (86). The highest number of participants were in the 22 - 25 years age group (36%) followed by the 18-21 years age group (33.7%) and 26-29 years age group (26.8%). Maximum number of participants were recently graduated–after 2020 AD (80%). Similarly, most of the participants had less than 2 years of clinical experience (75.6%). Most were PCL nurses (41.8%) followed by BSc. nursing graduates (38.4%). Only 20% of the participants had received formal training on proper handling of the medications. More than 40% of the participants claim they spend 12 hours or more time studying and updating themselves on clinical knowledge in any week.

**Table 1:**
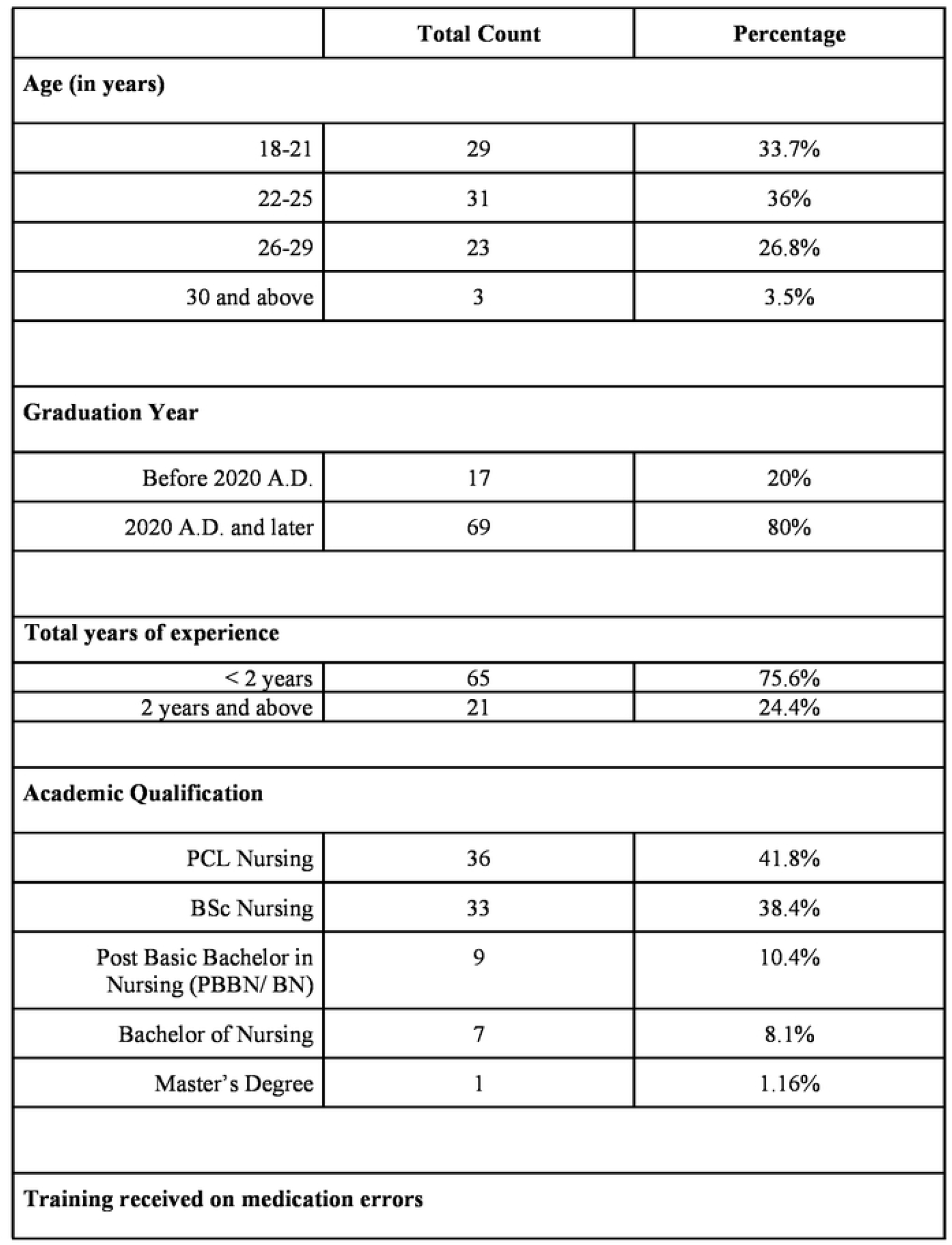
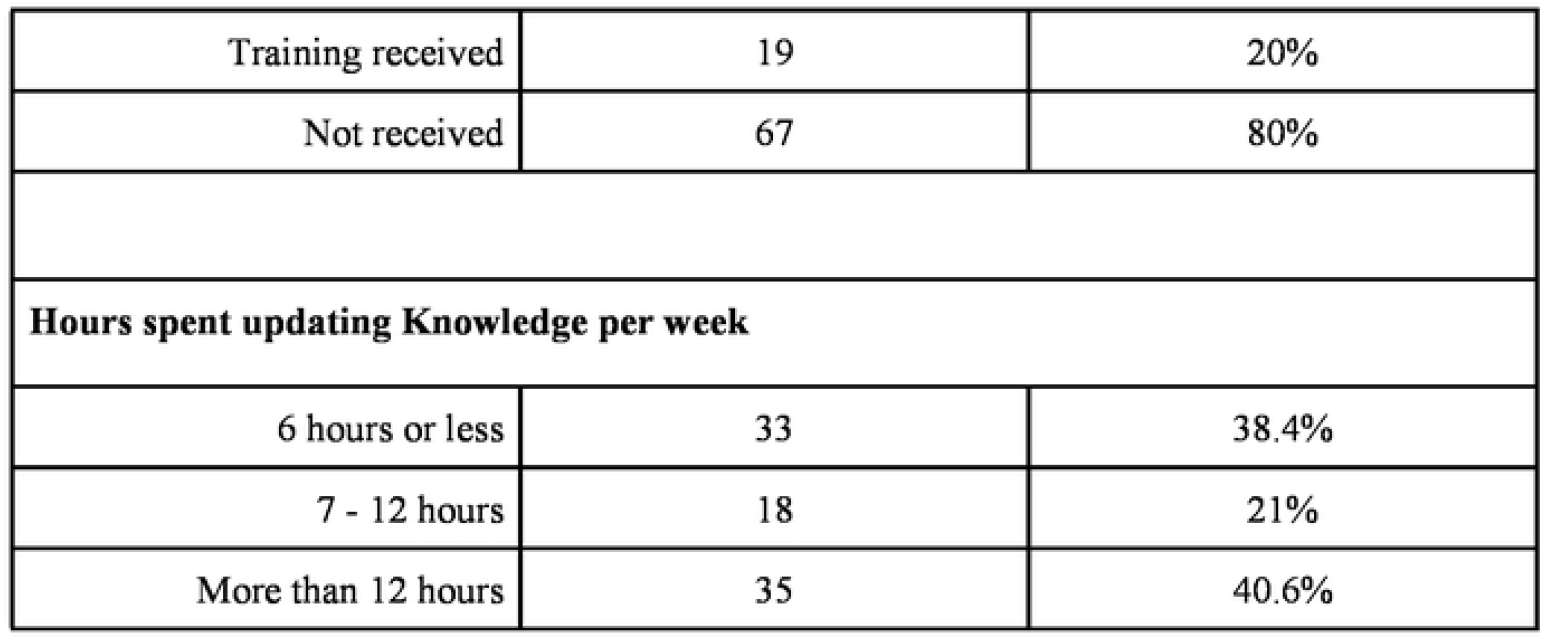
Demographic Characteristics of participants.

### 3.2 Knowledge

There was a significant increase in nurses’ knowledge after the education intervention in the use of CPOE reducing medication errors (p 0.00) and alarm noises and ward emergencies increase error risk (p 0.001). (Table 2)

**Table 2:** Knowledge-based assessment of nurses on Medication Error risks.

| Item | Pre-Test (N=70) |  | Post-Test (N=70) |  | Change from pre to post | McNemar's Test |  |  |
| --- | --- | --- | --- | --- | --- | --- | --- | --- |
|  | Agree | Disagree/ Neutral | Agree | Disagree/ Neutral |  | statistic | p-value (K2>K1) | p-value (if any difference) |
| Do you think dosage calculus of intravenous drugs reduces preparation errors? | 82.9% | 17.1% | 88.6% | 11.4% | 28.6% | 0.893 | 0.187 | 0.375 |
| Do you think use of Computerized drugs' prescription (CPOE) (computerized physician Order Entry system) reduces drug errors? | 42.9% | 57.1% | 82.9% | 17.1% | 51.4% | 5.582 | 0.000 | 0.000 |
| Do you think prepackaged package dispensation by pharmacy reduces medication error risk? | 64.3% | 35.7% | 74.3% | 25.7% | 44.3% | 1.263 | 0.106 | 0.211 |
| Do you think Informative protocols, posters, and brochures in wards promote reducing errors? | 92.9% | 7.1% | 97.1% | 2.9% | 7.1% | 1.350 | 0.091 | 0.182 |
| Do you think a pharmacist's assistance during drug preparation reduces error risk? | 71.4% | 28.6% | 80.0% | 20.0% | 34.3% | 1.229 | 0.112 | 0.223 |
| Do you think alarm noises and ward emergencies increase error risk? | 47.1% | 52.9% | 74.3% | 25.7% | 55.7% | 3.243 | 0.001 | 0.002 |
| Do you think Workload (double shifts, extra time) or frequent shift changes contribute to pharmacological therapy's errors? | 67.1% | 32.9% | 80.0% | 20.0% | 41.4% | 1.693 | 0.474 | 0.095 |

Significant differences could not be established in other areas of knowledge reducing error risk like IV drugs dosage calculus (p 0.187), prepackaged drug dispensation (p 0.106), use of informative protocols (p 0.091), importance of pharmacists’ assistance (0.112) and higher risk pertaining to higher workload (p 0.474).

### 3.3 Perception

Perceptions of the participants regarding various aspects of IV drug safety management before and after the intervention reveals a strong pre-existing consensus among participants on most safety practices An overwhelmingly high number of nurses agreed on the importance of error reporting, clinical skill evaluations, authoritative guidelines, and staff motivation that created a ceiling effect in the baseline survey. This provided a minimal space for measurable improvement through the intervention. The only statistically significant difference in participant’s perception was seen in effects of protocols/guidelines/procedures on ensuring proper management of therapeutic processes (p = 0.014). (Table 3)

**Table 3:** Perception of nurses on Medication Error reducing strategies.

| Variable | Pre-Test (N=70) |  | Post-Test (N=70) |  | Change from pre to post | McNemar's test |  |  |
| --- | --- | --- | --- | --- | --- | --- | --- | --- |
|  | Agree | Disagree/ Neutral | Agree | Disagree/ Neutral |  | Statistic | p-value (P2>P1) | p-value (if any difference) |
| Do you think ongoing and specific training on safe management of IV drugs could reduce the risk of error? | 97.1% | 2.9% | 94.3% | 5.7% | 8.57% | -0.815 | 0.791 | 0.418 |
| Do you think awareness for the prevention of error and clinical risk management could reduce the error during the preparation and administration's phases of drugs? | 90.0% | 10.0% | 94.3% | 5.7% | 12.86% | 1.000 | 0.160 | 0.321 |
| Do you think the worker's motivation can improve professional performance during the whole medication process? | 85.7% | 14.3% | 91.4% | 8.6% | 22.86% | 1.000 | 0.160 | 0.321 |
| Do you think, for secure management of the entire managing process of IV drugs, some authoritative guidelines drawn up taking into account the available scientific evidence are necessary? | 88.6% | 11.4% | 88.6% | 11.4% | 22.86% | 0.000 | 0.500 | 1.000 |
| Do you think protocols/guidelines/procedures can affect professional behavior, ensuring proper management of therapeutic processes? | 81.4% | 18.6% | 94.3% | 5.7% | 24.29% | 2.245 | 0.014 | 0.028 |
| Do you think clinical skills about safe management of drug therapy should be regularly evaluated? | 98.6% | 1.4% | 100.0 % | 0.0% | 1.43% | 1.000 | 0.160 | 0.321 |
| Do you think medication errors should be reported in order to become an opportunity for improving care? | 95.7% | 4.3% | 94.3% | 5.7% | 10.00% | -0.376 | 0.646 | 0.708 |

### 3.4 Behavior

The behavioral findings demonstrate near-perfect baseline compliance across all safety practices (98.6-100%), leaving minimal room for measurable improvement through intervention. While core protocol-driven behaviors like medication rights verification and infusion rate adherence showed unwavering 100% compliance - indicating thorough institutionalization - more variable practices like hand hygiene exhibited slight, non-significant declines. (Table 4)

**Table 4:** Behavior assessment of nurses on Medication Safety Practices.

|  | Pre-Test (N=70) |  | Post-Test (N=70) |  | Change from pre to post | McNemar's Test |  |  |
| --- | --- | --- | --- | --- | --- | --- | --- | --- |
| Variable | Agree | Disagree/ Neutral | Agree | Disagree/ Neutral |  | Statistic | p-value (B2>B1) | p-value (if any difference ) |
| Do you wash hands before the preparation and administration of therapy? | 98.6% | 1.4% | 91.4% | 8.6% | 10.00% | -1.926 | 0.971 | 0.058 |
| Do you check vital signs before and after administration of vasoactive drugs (Dopamine, Dobutamine, Adrenaline, Noradrenaline, Nitroglycerine, etc)? | 100.0% | 0.0% | 100.0% | 0.0% | 0.00% | . | . | . |
| Do you follow the rate of infusion of solutions for IV administration (such as antibiotics, chemotherapy, heparin, etc)? | 100.0% | 0.0% | 100.0% | 0.0% | 0.00% | . | . | . |
| Do you follow the ‘Rights’ of medications (Right prescription, Right drug, right patient, right dose, right route, etc)? | 100.0% | 0.0% | 100.0% | 0.0% | 0.00% | . | . | . |
| Do you perform a double check to verify the correct correspondence between prescription, preparation and administration of IV drugs, before administration? | 100.0% | 0.0% | 97.1% | 2.9% | 2.86% | -1.425 | 0.921 | 0.159 |

## 4. Discussion

We performed a quantitative, quasi-experimental study with a pretest-posttest design. This study assessed the effect of an educational training on the knowledge, attitudes, and practices (KAP) of the critical care nurses working in a tertiary level hospital concerning medication errors. The findings present a complex scenario, indicating notable improvement in specific knowledge and perception domains. Although behavioral change has remained relatively unchanged–perhaps due to elevated baseline adherence–which might be due to education and training imparted by nursing educators.

### Knowledge Domain

Educational interventions substantially enhanced nurses’ comprehension of the utilization of Computerized Physician Order Entry (CPOE) systems, thereby decreasing medication errors (p = 0.00). Kwiecień-Jaguś et al. emphasized that medication errors in ICU settings were substantially reduced by CPOE systems when used in conjunction with appropriate training, as they minimized transcription and dosage errors. ^8^ Piriou et al reiterated the importance of using computerized systems for drug prescription at different levels for eg. active checking measures like barcodes, use of programmable power syringes connected to a computer. ^15^ Similarly, Dionisi et al. underscored that nurses’ confidence and accuracy in medication administration were improved by structured education on CPOE systems. ^16^ The positive reception among nurses after the intervention indicates an openness to adopting such tools, especially when their practical benefits are clearly demonstrated. This intervention can be used to further explore the potential of targeted interventions in transforming perceptions of technology in healthcare. Johnson et al. observed that interruptions during medication preparation and administration were strongly linked to procedural failures and clinical errors.^17^ Our study demonstrated a significant association between alarm noises, ward emergencies, and increased error risk (p = 0.001). This finding highlights the nurses’ acknowledgment that continuous distracting alarm noises as well as frequent ward emergencies can impair focus, judgment and decision-making. This can lead to increased errors in care, including medication errors. This highlights the need for interventions that can mitigate these risks, like optimized alarm settings and improved communications during emergencies. Regular ward emergency exercises can improve both communication and situational awareness in nurses. In ICU settings, nurses frequently encounter distractions that jeopardize patient safety. In order to mitigate these risks, strategies such as the implementation of “quiet zones” and the reduction of interruptions during medication rounds have been recommended.^17^

Our findings indicate no significant improvement in knowledge related to IV drug dosage calculation (p = 0.187). This is concerning, as errors in IV medication administration are among the most frequent and severe in ICU settings. Westbrook et al. reported that incorrect IV rates and dosages accounted for a significant proportion of medication errors, emphasizing the need for continuous education and skill development^18^. Hamdan et al. also highlighted gaps in nurses’ training regarding IV drug preparation and administration.^19^ This may have been due to a high pretest score of 83% followed by 88.6% posttest score improvement. Still, we acknowledge that there is room for improvement. Our findings on prepackaged drug dispensation (p = 0.106) resonate with the challenges highlighted by Kwiecień-Jaguś et al., who noted that while prepackaged medications reduce manual errors, their effectiveness depends on proper labeling and staff familiarity.^8^ They recommended regular audits and pharmacist-led training to ensure safe implementation. Our study found no significant improvement in knowledge regarding the use of informative protocols (p = 0.091). Informatics tools, such as clinical decision support systems (CDSS), have been shown to reduce medication errors by providing real-time alerts and evidence-based recommendations.^15^ Dionisi et al. found that CDSS tools significantly reduced medication errors by providing real-time alerts and evidence-based guidelines. However, they emphasized that staff training is crucial for maximizing the utility of these systems.^16^ It is also closely aligned with one of the major objectives of our study and the findings show that we were able to correct nurses’ viewpoint on the importance of informative protocols by 10% posttest from 64%. Medication in critical care based training which was only obtained by 20% nurses can reflect this finding. We acknowledge that there is an emergent need for such training in our center. The lack of significant improvement in understanding the role of pharmacists (p = 0.112) highlights a missed opportunity to leverage their expertise. Escrivá Gracia et al. demonstrated that pharmacist-led interventions, such as medication reconciliation and error reporting, significantly reduced errors in ICU settings.^20^ Collaborative approaches between nurses and pharmacists were found to enhance patient safety. Our findings on the non-significant association between workload and error risk (p = 0.474) contrast with the broader literature. Dionisi et al. identified excessive workload as a critical factor contributing to medication errors, particularly in high-stress environments like ICUs.^16^ They suggested optimizing nurse-to-patient ratios and implementing time-management strategies to alleviate workload pressures.

### Perception Domain

The perception section revealed generally strong pre-existing beliefs regarding safe medication practices. However, one item stood out; there was a significant improvement in the belief that protocols/guidelines/procedures shape professional behavior and support the proper management of therapeutic processes (p = 0.014). This highlights the critical importance of structured frameworks in standardizing nursing practices and minimizing variability in medication management. Other studies have also clearly demonstrated that structured guidelines influence professional behavior, ensuring proper therapeutic management.^21^ To address this, we need to implement protocols specifically tailored to the Nepali critical care context, developed by the Nepalese Society of Critical Care Medicine (NSCCM). These protocols are applicable in both advanced facilities, and resource-limited settings. Additionally, essential drug information, including accurate dosages for inotropic and vasoactive medications, should be readily available in every ICU room, ensuring precise and reliable medication administration.

The remaining perception items showed little to no significant changes, likely due to the high levels of agreement in the baseline and limited potential for short-term shifts. For instance, 97% of participants already acknowledge that ongoing and specific training on IV drug safety helps minimize errors. However, this does not imply that no further improvements are necessary. Given the frequent introduction of new medications in critical care, continuous learning and training remain essential, as insufficient education on new drugs has previously been identified as a major factor contributing to errors.^21^ These findings should be interpreted cautiously, recognizing that improvement is always possible, as the process is continuously evolving.

### Behavior Domain

The behavioral findings demonstrate near-perfect baseline compliance across all safety practices (91.4-100%), leaving minimal room for measurable improvement through intervention. While core protocol-driven behaviors like medication rights verification and infusion rate adherence showed unwavering 100% compliance - indicating thorough institutionalization - more variable practices like hand hygiene exhibited slight, non-significant declines. These findings are better than other research done in similar settings. 100% compliance of nurses in checking vital signs during administration of vasoactive drugs compared to 88.8% in the research. Similarly, 98.8% nurses washed hands properly before medication administration compared to only 87.9% in the research. ^19^ The results highlight both the success of existing safety protocols in achieving high adherence for critical behaviors and the inherent limitations of educational interventions in enhancing already-optimized practices. These findings suggest future quality initiatives should focus on identifying and addressing specific performance gaps rather than broadly reinforcing well-established protocols, while maintaining vigilance to prevent erosion of existing high compliance standards. The data ultimately reveals an environment where fundamental medication safety behaviors have become deeply embedded in clinical routines.

## 5. Conclusion

This study highlights the significant impact of targeted education on improving critical care nurses’ understanding of computerized order entry and alarm-related risks, reinforcing protocol adherence where initial agreement was incomplete. Despite these improvements, the near-perfect baseline compliance with core medication safety behaviors and strong pre-existing safety perceptions indicate that most fundamental practices are already well institutionalized within the clinical setting. These findings suggest that in mature safety cultures, focused interventions are more effective than broad retraining efforts, as educational programs yield greater success when directed at specific areas requiring enhancement rather than reinforcing well-established norms. Future quality improvement initiatives should integrate targeted education on emerging risks with ongoing reinforcement of existing high standards to maintain optimal patient safety. This approach underscores a shift toward data-driven, customized strategies for advancing pharmaceutical safety in well-developed clinical environments.

## 6. Limitations of the study

This study acknowledges several constraints inherent to its cross-sectional survey design. Additionally, potential participant omission may limit sample representativeness. Also, as a quasi-experimental design lacking randomization, confounding variables (e.g., prior experience) could influence outcomes. This study was analyzed using McNemar’s test analysis, which was restricted to 70 participants, reducing statistical power and necessitating larger-scale validation. Furthermore, selection bias might be seen due to single-center recruitment. Lastly, social desirability bias may persist in self-reports despite anonymization. The absence of significant behavioral improvements may reflect either ceiling effects from high baseline adherence or the inherent limitations of self-reported versus observed behaviors, including recall bias. These factors collectively underscore the need for cautious interpretation of findings.

## Data Availability

All data produced in the present study are available upon reasonable request to the authors

## Acknowledgements

We would like to express our deepest gratitude to the nurses for their invaluable assistance in data collection and enthusiastic participation in this single-center ICU research. Their dedication and commitment to this research have been instrumental in its success. We are truly grateful for their contributions.

## List of Abbreviations

BN: Bachelor in Nursing
CDSS: Clinical decision support system
CPOE: Computerized physician order entry system
ICU: Intensive Care Unit
IV: Intravenous
HDU: High Dependency Unit
CCU: Coronary Care Unit
LMIC: Low and middle income country
NSCCM: Nepalese Society of Critical Care Medicine
p-value: Probability Value
RN: Registered Nurse

## References

1. Donchin Y, Gopher D, Olin M, et al. A look into the nature and causes of human errors in the intensive care unit. Crit Care Med. 1995;23(2):294–300. doi:10.1097/00003246-199502000-00015

2. Pronovost PJ, Thompson DA, Holzmueller CG, Lubomski LH, Morlock LL. Defining and measuring patient safety. Crit Care Clin. 2005;21(1):1–19. doi:10.1016/j.ccc.2004.07.006

3. Hussain E, Kao E. Medication safety and transfusion errors in the ICU and beyond. Crit Care Clin. 2005;21(1):91-110, ix. doi:10.1016/j.ccc.2004.08.003

4. Krähenbühl-Melcher A, Schlienger R, Lampert M, Haschke M, Drewe J, Krähenbühl S. Drug-Related Problems in Hospitals. Drug Saf. 2007;30(5):379–407. doi:10.2165/00002018-200730050-00003

5. Tissot E, Cornette C, Demoly P, Jacquet M, Barale F, Capellier G. Medication errors at the administration stage in an intensive care unit. Intensive Care Med. 1999;25(4):353–359. doi:10.1007/s001340050857

6. Camiré E, Moyen E, Stelfox HT. Medication errors in critical care: risk factors, prevention and disclosure. CMAJ Can Med Assoc J J Assoc Medicale Can. 2009;180(9):936–943. doi:10.1503/cmaj.080869

7. Moyen E, Camiré E, Stelfox HT. Clinical review: Medication errors in critical care. Crit Care. 2008;12(2):208. doi:10.1186/cc6813

8. Kwiecień-Jaguś K, Medrzycka-Dabrowska W, Kopec M. Understanding Medication Errors in Intensive Care Settings and Operating Rooms—A Systematic Review. Medicina (Mex). 2025;61(3):369. doi:10.3390/medicina61030369

9. Larsen GY, Parker HB, Cash J, O’Connell M, Grant MC. Standard drug concentrations and smart-pump technology reduce continuous-medication-infusion errors in pediatric patients. Pediatrics. 2005;116(1):e21–25. doi:10.1542/peds.2004-2452

10. Bullock J, Jordan D, Gawlinski A, Henneman EA. Standardizing IV Infusion Medication Concentrations to Reduce Variability in Medication Errors. Crit Care Nurs Clin North Am. 2006;18(4):515–521. doi:10.1016/j.ccell.2006.08.008

11. Shamliyan TA, Duval S, Du J, Kane RL. Just What the Doctor Ordered. Review of the Evidence of the Impact of Computerized Physician Order Entry System on Medication Errors. Health Serv Res. 2008;43(1p1):32-53. doi:10.1111/j.1475-6773.2007.00751.x

12. Pronovost P, Weast B, Schwarz M, et al. Medication reconciliation: a practical tool to reduce the risk of medication errors. J Crit Care. 2003;18(4):201–205. doi:10.1016/j.jcrc.2003.10.001

13. Cummings J, Bush P, Smith D, Matuszewski K. Bar-coding medication administration overview and consensus recommendations. Am J Health Syst Pharm. 2005;62(24):2626–2629. doi:10.2146/ajhp050222

14. Di Muzio M, Tartaglini D, De Vito C, La Torre G. Validation of a questionnaire for ICU nurses to assess knowledge, attitudes and behaviours towards medication errors. Ann Ig Med Prev E Comunita. 2016;28(2):113–121. doi:10.7416/ai.2016.2090

15. Piriou V, Theissen A, Arzalier-Daret S, et al. Preventing medication errors in anesthesia and critical care (abbreviated version). Anaesth Crit Care Pain Med. 2017;36(4):253–258. doi:10.1016/j.accpm.2017.04.002

16. Dionisi S, Giannetta N, Liquori G, et al. Medication Errors in Intensive Care Units: An Umbrella Review of Control Measures. Healthcare. 2022;10(7):1221. doi:10.3390/healthcare10071221

17. Johnson M, Sanchez P, Langdon R, et al. The impact of interruptions on medication errors in hospitals: an observational study of nurses. J Nurs Manag. 2017;25(7):498–507. doi:10.1111/jonm.12486

18. Westbrook JI, Rob MI, Woods A, Parry D. Errors in the administration of intravenous medications in hospital and the role of correct procedures and nurse experience. BMJ Qual Saf. 2011;20(12):1027–1034. doi:10.1136/bmjqs-2011-000089

19. Hamdan KM, Albqoor MA, Shaheen AM. Intravenous Medication Errors Among ICU Nurses: Differences In Knowledge Attitudes And Behavior. Open Nurs J. 2022;16(1):e187443462206201. doi:10.2174/18744346-v16-e2206201

20. Escrivá Gracia J, Brage Serrano R, Fernández Garrido J. Medication errors and drug knowledge gaps among critical-care nurses: a mixed multi-method study. BMC Health Serv Res. 2019;19(1):640. doi:10.1186/s12913-019-4481-7

21. Mahran DS. Patient Safety Culture and Application of Medication Safety Rules as Perceived by Nurses. Am J Nurs Sci. Published online January 1, 2016. Accessed May 7, 2025. https://www.academia.edu/59888311/Patient_Safety_Culture_and_Application_of_Medication_Safety_Rules_as_Perceived_by_Nurses

